# Cardiorenal effects of dual blockade with Angiotensin-converting enzyme inhibitors and Angiotensin receptor blockers in people with CKD: analysis of routinely collected data with emulation of a reference trial (ONTARGET)

**DOI:** 10.1101/2024.05.28.24307859

**Authors:** Paris J Baptiste, Angel YS Wong, Anna Schultze, Catherine M Clase, Clémence Leyrat, Elizabeth Williamson, Emma Powell, Johannes FE Mann, Marianne Cunnington, Koon Teo, Shrikant I Bangdiwala, Peggy Gao, Kevin Wing, Laurie Tomlinson

## Abstract

We aimed to explore whether the ONTARGET trial results, which led to an end of recommendations of dual angiotensin-converting enzyme inhibitor (ACEi) and angiotensin receptor blocker (ARB) use, extended to patients with chronic kidney disease (CKD) who were underrepresented in the trial.

We selected people prescribed an ACEi and/or an ARB in the UK Clinical Practice Research Datalink Aurum during 1/1/2001-31/7/2019. We specified an operational definition of dual users and applied ONTARGET eligibility criteria. We used propensity-score—weighted Cox-proportional hazards models to compare dual therapy to ACEi for the primary composite trial outcome (cardiovascular death, myocardial infarction, stroke, or hospitalisation for heart failure), as well as a primary composite renal outcome (≥50% reduction in GFR or end-stage kidney disease), and other secondary outcomes, including hyperkalaemia. Conditional on successfully benchmarking results against the ONTARGET trial, we explored treatment effect heterogeneity by CKD at baseline.

In the propensity-score—weighted trial-eligible analysis cohort (n=412 406), for dual therapy vs ACEi we observed hazard ratio (HR) 0.98 (95% CI: 0.93, 1.03), for the primary composite outcome, consistent with the trial results (ONTARGET HR 0.99, 95% CI: 0.92, 1.07). Dual therapy use was associated with an increased risk of the primary renal composite outcome, HR 1.25 (95% CI: 1.15, 1.36) vs ONTARGET HR 1.24 (1.01, 1.51) and hyperkalaemia, HR 1.15 (95% CI: 1.09, 1.22) in the trial eligible cohort, consistent with ONTARGET. The presence of CKD at baseline had minimal impact on results.

**Translational statement:** We extended ONTARGET trial findings of the comparative effectiveness of dual ARB and ACEi therapy use compared to ACEi alone for a composite cardiovascular outcome to UK patients at high-risk of cardiovascular disease, including those with CKD. As in ONTARGET, we found an increased risk of a composite renal outcome (≥50% reduction in GFR or end-stage kidney disease) and an increased risk of hyperkalaemia among dual users compared to ACEi alone. Consistent results were observed among patients with CKD at baseline. This is evidence against the hypothesis that dual blockade provides cardiorenal benefits among high-risk cardiovascular patients with CKD.

## INTRODUCTION

ONTARGET was a large global trial which compared the effects of a combination of ramipril (angiotensin-converting enzyme inhibitor (ACEi)) and telmisartan (angiotensin receptor blocker (ARB)) vs ramipril alone in patients at high-risk of cardiovascular events.^1^ Results from the trial, in conjunction with the ALTITUDE and VA-Nephron-D study,^2, 3^ changed practice, leading to an end of recommendations for dual ACEi and ARB therapy in patients with kidney disease.^4^ Despite these results there is uncertainty about whether dual blockade of the renin-angiotensin system could be effective at reducing adverse renal outcomes in patients with chronic kidney disease (CKD).^5, 6^

Reference trial emulation is a specific type of target trial emulation which can be used to add confidence to findings from observational comparative effectiveness studies.^7–10^ Whereas the majority of target trial emulation designs are based upon hypothetical RCTs,^11^ reference trial emulation involves emulation of an existing randomised controlled trial (RCT) to (1) inform observational study design and (2) benchmark results against.^12–14^ However, evidence on whether more complex interventions, such as dual therapy treatment arms, can be emulated in observational data is limited.

This study aimed to (1) emulate the ONTARGET trial in UK routinely collected healthcare data (2) benchmark the emulation analysis results with those of ONTARGET for the comparison of dual therapy vs ACEi alone and (3) extend analysis to investigate treatment heterogeneity by CKD status at baseline (conditional upon successful emulation of ONTARGET).

## METHODS

### The reference trial (ONTARGET)

The ONTARGET trial assessed the whether a combination of telmisartan (ARB) and ramipril (ACEi) was superior to ramipril (ACEi) alone and included 17 078 patients of whom 8502 were randomised to receive dual therapy. 23.4% of patients had CKD at baseline and mean creatinine was in the normal range at 93.8 μmol/l. Participants with coronary artery disease, peripheral artery disease, cerebrovascular disease or high-risk diabetes were included and those with heart failure were excluded. The primary outcome of the trial was a composite of cardiovascular-related death, myocardial infarction (MI), stroke or hospitalisation for heart failure. The study also investigated renal outcomes which included a primary composite of dialysis, doubling of creatinine or death, and individual components.

In ONTARGET, for the primary composite outcome there was no evidence of superiority of dual therapy compared with ACEi alone (HR of 0.99 (95% CI: 0.92, 1.07)), with evidence of an increase of adverse events, including hyperkalaemia, in participants treated with dual therapy.^15^ The HR was 1.09 (95% CI: 1.01, 1.18) for the primary composite renal outcome and there was no evidence of treatment effect heterogeneity by CKD status for this outcome (*P_int_*=0.80).

### Reference trial emulation using observational data

Briefly, we selected patients who were ever prescribed an ACEi and/or an ARB from 1/1/2001-31/7/2019 from primary care data. We then defined dual therapy and ACEi single therapy exposure periods before applying trial criteria to generate trial eligible periods. Methods are detailed in a previously published protocol and summarised in Supplementary Figure S1.^16^ Key design aspects of the ONTARGET trial and this emulation are presented in Supplementary Table S1. The steps involved in the creation of the dataset are further described below.

#### Data sources

We selected patients from the UK Clinical Practice Research Datalink (CPRD) Aurum. As of 2021, CPRD Aurum included 13 million alive patients currently registered at a contributing general practice. This represents ∼20% of the UK population.^17^ Patients were required to have been registered at an up-to-standard practice (ensuring adequate data quality) in CPRD for at least 12 months at the time of their first selected prescription. Only those patients who had linked hospitalisation data from Hospital Episode Statistics (HES) and death registrations from the Office for National Statistics (ONS) were included.^18, 19^

#### Outcomes

We compared primary and renal outcomes aligned with the clinical trial between dual (ARB and ACEi) users vs ACEi alone:^1, 20^

Cardiovascular outcomes:

- Primary outcome: composite of cardiovascular death, myocardial infarction (MI), stroke or hospital admission for congestive heart failure
- Main secondary outcome: composite of cardiovascular death, MI, or stroke

Renal outcomes:

- Primary renal outcome: composite of loss of glomerular filtration rate (GFR) (defined as: 50% reduction in estimated GFR (eGFR)) or development of end-stage kidney disease (ESKD) (defined as: start of kidney replacement therapy (KRT) or development of eGFR < 15ml/min/1.73m^2^).
- ≥50% reduction in eGFR
- ESKD (defined as: start of KRT or development of eGFR < 15ml/min/1.73m^2^)
- Doubling of serum creatinine

Safety outcome:

- Hyperkalaemia (potassium > 5.5mmol/l)

Renal outcomes studied in ONTARGET included both prespecified outcomes and adverse events, some of which were defined by the lead clinical investigators locally.^1^ We chose a definition of loss of kidney function that includes loss of 50% GFR rather than doubling of creatinine, in keeping with current methodological practice.

GFR was calculated using the CKD-Epi equation 2009 without reference to ethnicity.^21^

#### Treatment strategies

##### Single therapy exposure

Prescriptions for an ACEi with <90 days between the calculated end date and start of subsequent prescription were combined to create exposed periods. If a patient stopped and restarted treatment, they could have multiple exposed periods. Therefore, a patient could contribute multiple eligible exposed periods, and as in a trial, a patient could meet the trial eligibility criteria on more than one occasion.^22^

##### Dual therapy exposure

We defined dual therapy users as patients with overlapping prescriptions of an ACEi and ARB who had a subsequent prescription for the 1^st^ agent within 90 days of the duration of the 2^nd^ prescription for the 2^nd^ agent. Follow-up was then started from the date this operational definition was met, i.e., the date of the 2^nd^ prescription for the 1^st^ agent and eligibility criteria was assessed at this time point (Figure 1). Many patients switch between an ACEi and ARB during their treatment history. Therefore, we required patients to have a 2^nd^ prescription for the 1^st^ agent after the 2^nd^ agent was added to ensure we identified dual users rather than people switching treatment. Including only those who met this operational definition could introduce bias by excluding those who die or who have early adverse events, so impact of alternate definitions were also assessed in sensitivity analyses.

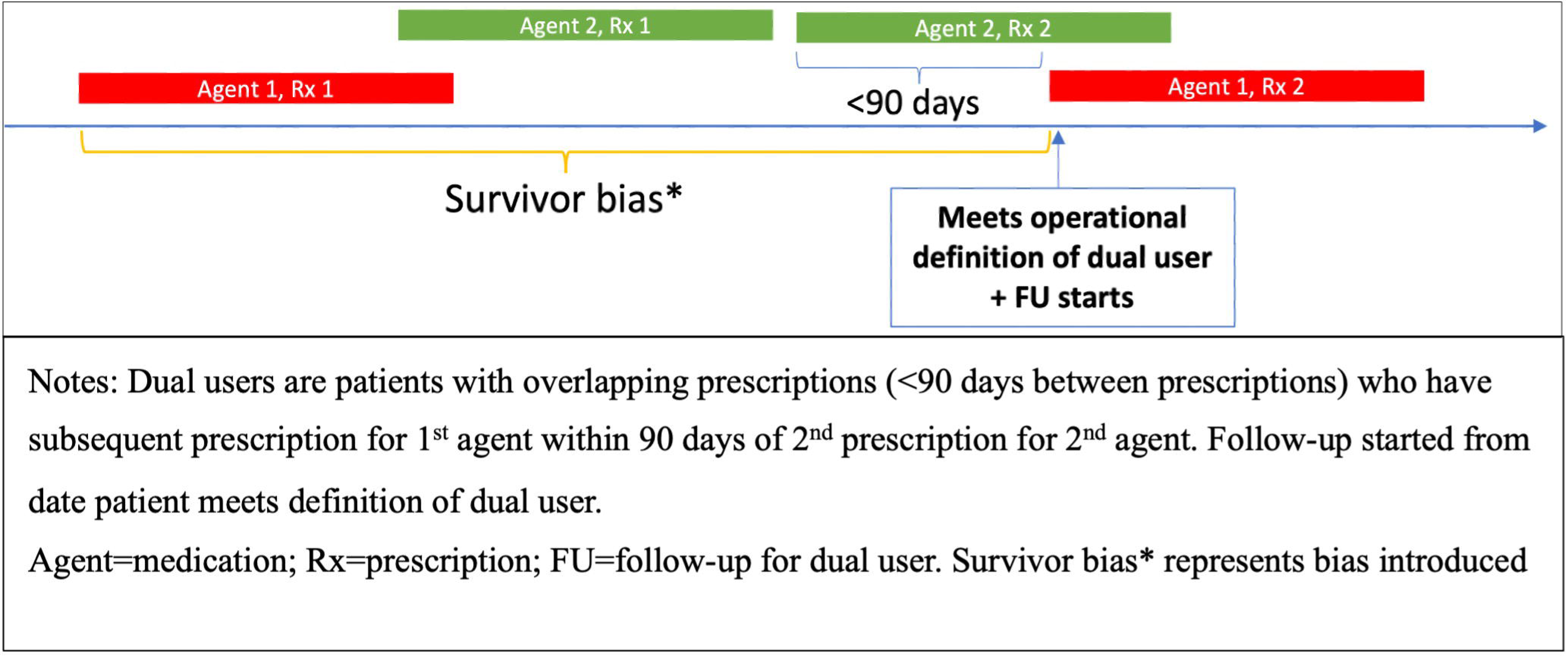

Eligibility criteria were the same as the ONTARGET trial and assessed at the start of each exposed period,^16^ generating trial-eligible periods. ONTARGET trial criteria and the interpretation in CPRD is detailed in the Supplementary Table S2-S3.

### Statistical analysis

#### Achieving balance across exposure groups

To preserve sample size, we used propensity-score—weighting, rather than matching, to achieve balance among exposure groups. The propensity score was estimated using a logistic regression model for the probability of receiving an ACEi.^23^ As with previous work,^14^ we selected one random trial-eligible period per patient in the trial-eligible ACEi and dual therapy exposure groups. A random period was chosen as opposed to the first period as selecting the first period may bias results to new users.

Variables considered in the propensity-score model were chosen based on a-priori knowledge and included baseline demographics, socio-economic status, medication, and clinical history. To account for the potential time-related bias introduced by changing usage of dual blockade over time as a result of published trials and European Medicines Agency guidance,^24^ we included time since first trial eligible period in our propensity-score model.^25^ Included variables are displayed in Supplementary Table S4.

#### Estimating treatment effects

Treatment effectiveness was assessed using a time-to-event analysis weighted by propensity-score, using robust standard errors in a Cox proportional hazards model. Patients were followed until the earliest of outcome, death, transferred out of practice, practice last collection or a maximum of 5.5 years of follow up. Treatment cessation of one or both agents during follow-up were ignored, to match the intention-to-treat approach from the reference trial.

#### Benchmarking

We pre-specified criteria to confirm successful benchmarking against the ONTARGET trial primary composite outcome.^16^ This included if the HR estimates from the observational study for dual therapy compared to ACEi were between 0.9-1.12 and the 95% CI for the HR contained 1.

Formal criteria were not used to assess similarity of findings compared to ONTARGET results for secondary and renal outcomes, as these differed to those studied in ONTARGET.

#### Extending analysis to trial- underrepresented group of those with CKD

Conditional on the validation criteria being met, we examined whether there was evidence to suggest a benefit of dual blockade among patients with CKD in routine care. In this study we defined CKD as eGFR<60ml/min/1.73m^2^. Aided by a larger sample size with more diverse characteristics than the ONTARGET trial and therefore a higher proportion of patients with CKD, we had greater power than the trial to detect treatment effect heterogeneity. This was assessed by fitting an interaction term between CKD at baseline and treatment in the propensity-score—weighted Cox model. The balance of characteristics was assessed within stratum of CKD at baseline using standardised mean differences.

#### Sensitivity analyses

We assessed the impact of bias due to the definition of dual user chosen by benchmarking results for the two alternate definitions of a dual user against the ONTARGET trial. Details about the two alternate dual user definitions are provided in the Appendix and displayed graphically in Supplementary Figure S2.

We included a sensitivity analysis to assess the robustness of a complete-case analysis to departures from a missing completely at random assumption, for variables included in our propensity-score model that had >10% missing data. We used multiple imputation with chained equations to re-estimate treatment effects for the primary cardiovascular outcome for the benchmarking analysis using inverse probability weighting.^26–28^

Because of likely differences in baseline proteinuria with patients prescribed dual therapy or single therapy and to explore heterogeneity by proteinuria status, we also included a sensitivity analysis stratifying analysis by proteinuria status at baseline, defining proteinuria as albumin-creatinine ratio >3.

Finally, to explore if potential differences in duration of prior antihypertensive exposure biased results, we repeated analyses for the primary outcome adjusting for time since first exposure to an ARB, ACEi or dual therapy.

## RESULTS

### Baseline characteristics

Among the 2,195,718 patients who had ACEi or dual therapy exposure periods, 412,402 met the trial criteria and were included in the propensity-score—weighted analysis cohort. Prior to propensity-score weighting, patients receiving dual therapy treatment were more likely to have higher baseline blood pressure, higher creatinine and be from Black or South Asian ethnic groups compared to patients prescribed an ACEi alone (Table 1). More patients receiving dual therapy had diabetes with proteinuria compared with patients receiving ACEi alone, 19.8% vs 11.9% in the dual therapy and ACEi exposure groups, respectively. Characteristics were balanced after weighting and are displayed in Supplementary Table S5. The analysis cohort included 392 475 ACEi and 19 931 dual therapy patients (Figure 2).

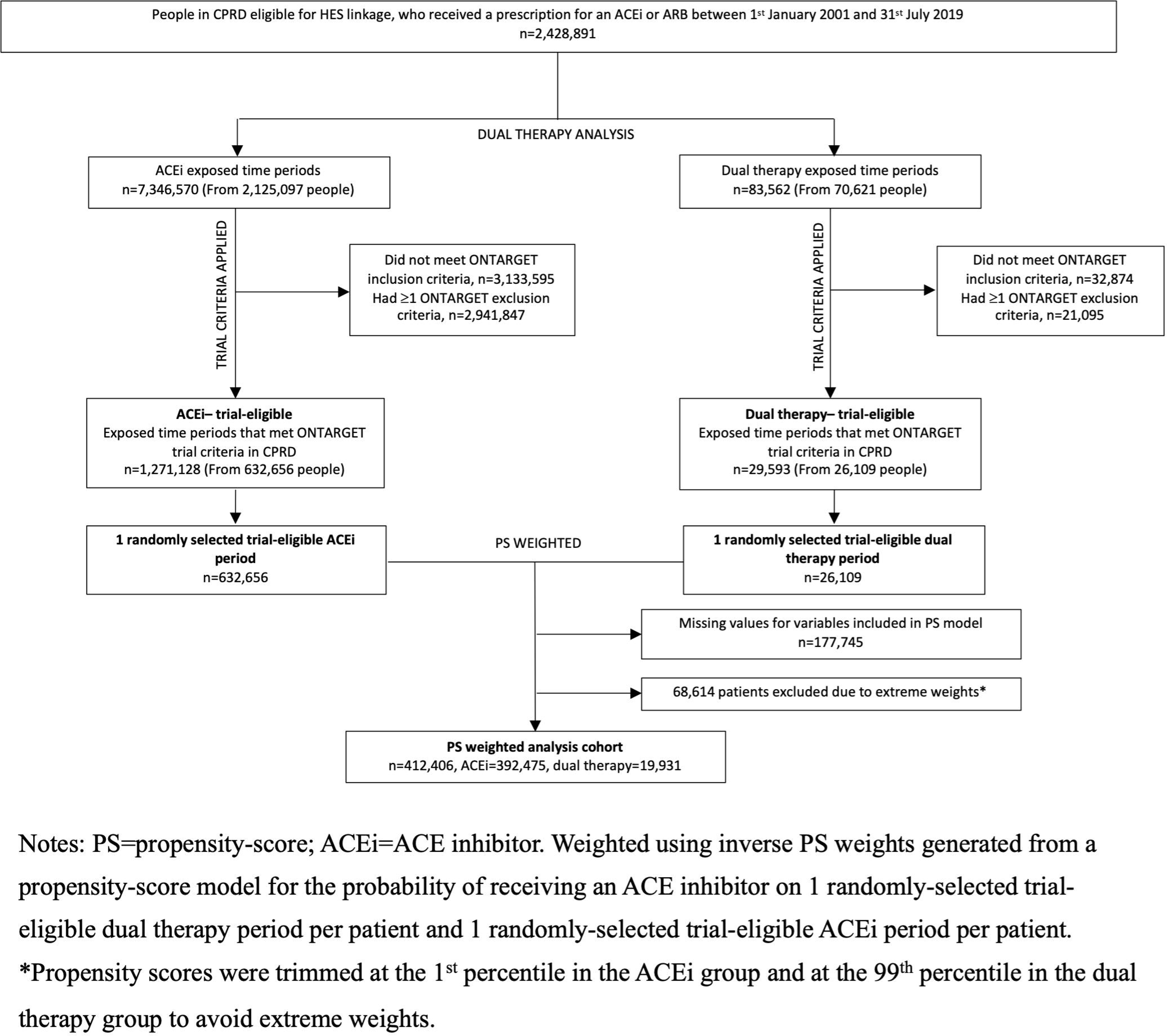

**Table 1.** Baseline characteristics of trial-eligible patients included in analysis compared to ONTARGET.

| Characteristic | Dual therapy<br>N=19 931 | ACEi<br>N=392 475 | ONTARGET<br>N=17 078 |
| --- | --- | --- | --- |
| Age (year) – mean (SD) | 70.4 (8.8) | 70.5 (9.4) | 66.4 (7.2) |
| Systolic BP (mmHg) – mean (SD) | 147.7 (20.0) | 144.3 (19.8) | 141.9 (17.5) |
| Diastolic BP (mmHg) – mean (SD) | 79.7 (10.8) | 79.4 (10.9) | 82.1 (10.4) |
| Body-mass index – mean (SD) | 29.7 (5.9) | 28.9 (5.8) | 28.1 (4.7) |
| Creatinine (μmol/l) – mean (SD) | 96.4 (28.9) | 92.8 (26.8) | 94.1 (24.3) |
| Potassium (mmol/l) – mean (SD) | 4.5 (0.5) | 4.4 (0.5) | 4.4 (0.4) |
| Female sex – no. (%) | 10388 (52.1) | 186817 (47.6) | 4581 (26.8) |
| Ethnic group – no. (%) |  |  |  |
| Black | 851 (4.3) | 10888 (2.8) | 414 (2.4) |

| **Characteristic** | **Dual therapy**<br>N=19 931 | **ACEi**<br>N=392 475 | **ONTARGET**<br>N=17 078 |
| --- | --- | --- | --- |
| Other | 329 (1.7) | 5233 (1.3) | 3233 (19.0) |
| South Asian | 1565 (7.9) | 19152 (4.9) | 921 (5.4) |
| Missing | - | - | 4 (<0.1) |
| White | 17186 (86.2) | 357202 (91.0) | 12495 (73.2) |
| **Clinical history – no. (%)** |  |  |  |
| CAD<sup>a</sup> | 12296 (61.7) | 275353 (70.2) | 12735 (74.6) |
| Cerebrovascular disease<sup>b</sup> | 2149 (10.8) | 41967 (10.7) | 3584 (21.0) |
| PAD<sup>c</sup> | 1930 (9.7) | 36542 (9.3) | 2307 (13.5) |
| Diabetes | 13462 (67.5) | 237841 (60.6) | 6366 (37.3) |
| High-risk diabetes<sup>d</sup> | 12574 (63.1) | 189023 (48.2) | 4706 (27.6) |
| **Smoking status – no. (%)** |  |  |  |
| Non-smoker | 6040 (30.3) | 106185 (27.1) | 6083 (35.6) |
| Current smoker | 4860 (24.4) | 105931 (27.0) | 2163 (12.7) |
| Past smoker | 9031 (45.3) | 180359 (46.0) | 8808 (51.6) |
| Missing | - | - | 24 (0.1) |
| **Alcohol status – no. (%)** |  |  |  |
| Non-drinker | 3711 (18.6) | 61706 (15.7) | 10172 (59.6) |
| Current drinker | 11578 (58.1) | 245276 (62.5) | 6897 (40.4) |
| Past drinker | 2647 (13.3) | 49349 (12.6) | - |
| Missing | 1995 (10.0) | 36144 (9.2) | 9 (0.1) |
| **Medication<sup>e</sup> – no. (%)** |  |  |  |
| Alpha-blocker | 2817 (14.1) | 35819 (9.1) | 745 (4.4) |
| Oral anticoagulant agent | 1147 (5.8) | 27971 (7.1) | 1296 (7.6) |
| Antiplatelet agent | 1442 (7.2) | 31143 (7.9) | 1858 (10.9) |
| Aspirin | 7809 (39.2) | 135630 (34.6) | 12934 (75.7) |
| Beta-blocker | 6492 (32.6) | 120585 (30.7) | 9723 (56.9) |
| Calcium-channel blocker | 7917 (39.7) | 130355 (33.2) | 5685 (33.3) |
| Digoxin | 557 (2.8) | 13976 (3.6) | 555 (3.3) |
| Diuretics | 9519 (47.8) | 150598 (38.4) | 4805 (28.1) |
| Diabetic treatment | 7077 (35.5) | 101350 (25.8) | 5334 (31.2) |
| Nitrates | 1730 (8.7) | 36904 (9.4) | 4987 (29.2) |
| Statins | 11508 (57.7) | 210608 (53.7) | 10489 (61.4) |
| N= number of patients; SD=standard deviation; no. (%)=number (percent); BP= blood pressure;<br>CAD=coronary artery disease; PAD=peripheral artery disease; CKD=chronic kidney disease<br>(eGFR<60ml/min/1.73m<sup>2</sup>)<br>One third of ONTARGET participants received both ramipril plus telmisartan. |  |  |  |

| Characteristic | Dual therapy<br>N=19 931 | ACEi<br>N=392 475 | ONTARGET<br>N=17 078 |
| --- | --- | --- | --- |
| <sup>a</sup> Includes diagnosis of: MI at least 2 days prior, angina at least 30 days prior, angioplasty at least 30 days prior, CABG at least 4 years prior<br><sup>b</sup> Includes diagnosis of: stroke/TIA<br><sup>c</sup> Includes diagnosis of: limb bypass surgery, limb/foot amputation, intermittent claudication<br><sup>d</sup> Includes DM with: retinopathy, neuropathy, chronic kidney disease, proteinuria or other complication<br><sup>e</sup> Within 3 months prior to eligible start date. Antiplatelet agent= clopidogrel/ticlopidine.<br>In the categorisation of ethnicity in ONTARGET South Asian ethnic group included Other Asian and Black included Black African and Colored African as described in the trial CRF. |  |  |  |

### Benchmarking results

#### Primary outcome

The primary composite cardiovascular outcome occurred in 3478 (17.5%) patients in the dual therapy group and 69 258 (17.7%) patients in the ACEi group. Over a 5.5 year follow up incidence rates were 4.02 and 4.12 per 100 person-years in the in the dual therapy and ACEi groups, respectively. The risk of the primary cardiovascular outcome was similar among dual therapy and ACEi users, HR 0.98 (95% CI: 0.93, 1.03), consistent with the ONTARGET result (HR 0.99 (95% CI: 0.92, 1.07)) and met the pre-defined criteria for successful benchmarking (Table 2).

**Table 2.** Number of events and results for the cardiovascular, renal and safety outcomes for dual therapy vs ACEi using a propensity-score—weighted analysis of trial-eligible patients in CPRD Aurum compared to ONTARGET.

| Outcome | CPRD |  |  | ONTARGET |
| --- | --- | --- | --- | --- |
|  | Dual therapy<br>(N=19 931) | ACEi<br>(N=392 475) | Dual therapy vs<br>ACEi | Dual therapy vs<br>ramipril |

|  |  |  | (N=412 406) | (N=17 078) |
| --- | --- | --- | --- | --- |
|  | *Number (percent)* |  | *Hazard ratio (95% CI)* |  |
| Primary composite | 3748 (17.5) | 69258 (17.7) | 0.98 (0.93, 1.03) | 0.99 (0.92, 1.07) |
| Main secondary outcome | 2853 (14.3) | 57220 (14.6) | 1.01 (0.95, 1.07) | 1.00 (0.93, 1.09) |
| Primary renal outcome<sup>1</sup> | 1545 (8.5) | 19271 (5.6) | 1.25 (1.15, 1.36) | 1.24 (1.01, 1.51)<sup>2</sup> |
| 50% reduction in GFR<sup>1</sup> | 1468 (8.0) | 17788 (5.2) | 1.23 (1.13, 1.34) |  |
| ESKD | 578 (2.9) | 6733 (1.7) | 1.38 (1.19, 1.60) |  |
| Doubling of creatinine<sup>1</sup> | 939 (5.1) | 11186 (3.3) | 1.20 (1.07, 1.34) | 1.20 (0.96, 1.50) |
| Hyperkalaemia | 2922 (14.7) | 42020 (10.7) | 1.15 (1.09, 1.22) | 1.58 (1.26, 1.99)<sup>3</sup> |
| GFR: glomerular filtration rate; ESKD: end-stage kidney disease.<br>Primary composite outcome: death from cardiovascular causes, myocardial infarction, stroke, or hospitalisation for heart failure.<br>Main secondary outcome: death from cardiovascular causes, myocardial infarction, or stroke.<br>Primary renal outcome: composite of loss of GFR ($\geq 50\%$ reduction in GFR) or development of end-stage kidney disease (GFR$<15$ or start of kidney replacement therapy).<br>ESKD: GFR$<15$ or start of kidney replacement therapy<br>CPRD weighted analysis includes 1 randomly-selected trial-eligible period per patient. Propensity-score—weighted with robust standard errors.<br><sup>1</sup>Outcome studied among patients in CPRD with non-missing creatinine at baseline (dual: n=18 270; ACEi: n=343 983).<br>ONTARGET results are from published findings.<br><sup>2</sup>ONTARGET studied a composite renal outcome of dialysis or doubling of serum creatinine which differed from our primary renal composite outcome, which is dialysis or $\geq 50\%$ reduction in GFR, so results are not directly comparable.<br><sup>3</sup>ONTARGET studied hyperkalaemia at 6 weeks therefore not directly comparable to our results which studied hyperkalaemia during total follow up. |  |  |  |  |

#### Renal and safety outcomes

Results were consistent with ONTARGET for the main secondary cardiovascular outcome (cardiovascular death, MI or stroke) with HR of 1.01 (95% CI: 0.95, 1.07) vs HR 1.00 (95% CI 0.93, 1.09) for ONTARGET (Table 2). For the primary renal composite outcome (loss of GFR or development of ESKD), risk was increased among dual therapy users compared with users of ACEi alone with propensity-score—weighted HR of 1.25 (95% CI: 1.15, 1.36). This was consistent with the ONTARGET composite outcome of dialysis or doubling of creatinine (ONTARGET HR 1.24 (1.01, 1.51)). Similar results were observed when the individual components of the composite renal outcome were assessed. For doubling of creatinine the HR was also consistent with the trial findings, HR 1.20 (95% CI: 1.07, 1.34), ONTARGET HR 1.20 (95% CI: 0.96, 1.50) (Table 2).^1^ Consistent with ONTARGET reported conclusions, dual therapy was associated with an increased risk of hyperkalaemia compared with ACEi alone, HR 1.15 (95% CI: 1.09, 1.22).

### Extending the analysis to trial-underrepresented group of those with CKD

Among those with non-missing baseline CKD status (87.8%), 6816 (37.3%) of patients had CKD in the dual therapy group and 107 371 (31.2%) patients had CKD in the ACEi group. Among those who had CKD at baseline, the primary outcome occurred in 1593 (23.4%) patients in the dual therapy group and 27 579 (25.7%) patients in the ACEi group. Among those who did not have CKD at baseline, the number of events for the primary composite outcome was 1585 (13.8%) in the dual therapy group and 32 328 (13.7%) in the ACEi group. There was no evidence of treatment effect heterogeneity by CKD status for the primary composite cardiovascular outcome (*P_int_* = 0.14). For dual therapy vs ACEi use, the HRs were 0.95 (95% CI: 0.87, 1.02) and 1.03 (95% CI: 0.95, 1.11) among those with and without CKD at baseline, respectively (Figure 3). Results were similar for the main secondary outcome (*P_int_* = 0.62). Dual therapy use was associated with an increased risk of the primary renal composite outcome, with no evidence of treatment heterogeneity by CKD status (*P_int_* = 0.92). For dual therapy vs ACEi, HRs were 1.26 (95% CI: 1.12, 1.41) and 1.25 (95% CI: 1.10, 1.42) among those with and without CKD at baseline, respectively (Figure 3). Similar results were observed for other renal outcomes (*P_int_* = 0.43 to 0.96). Dual use was associated with an increased risk of hyperkalaemia and there was no evidence of heterogeneity by CKD status (*P_in_*_t_ = 0.24) (Figure 3).

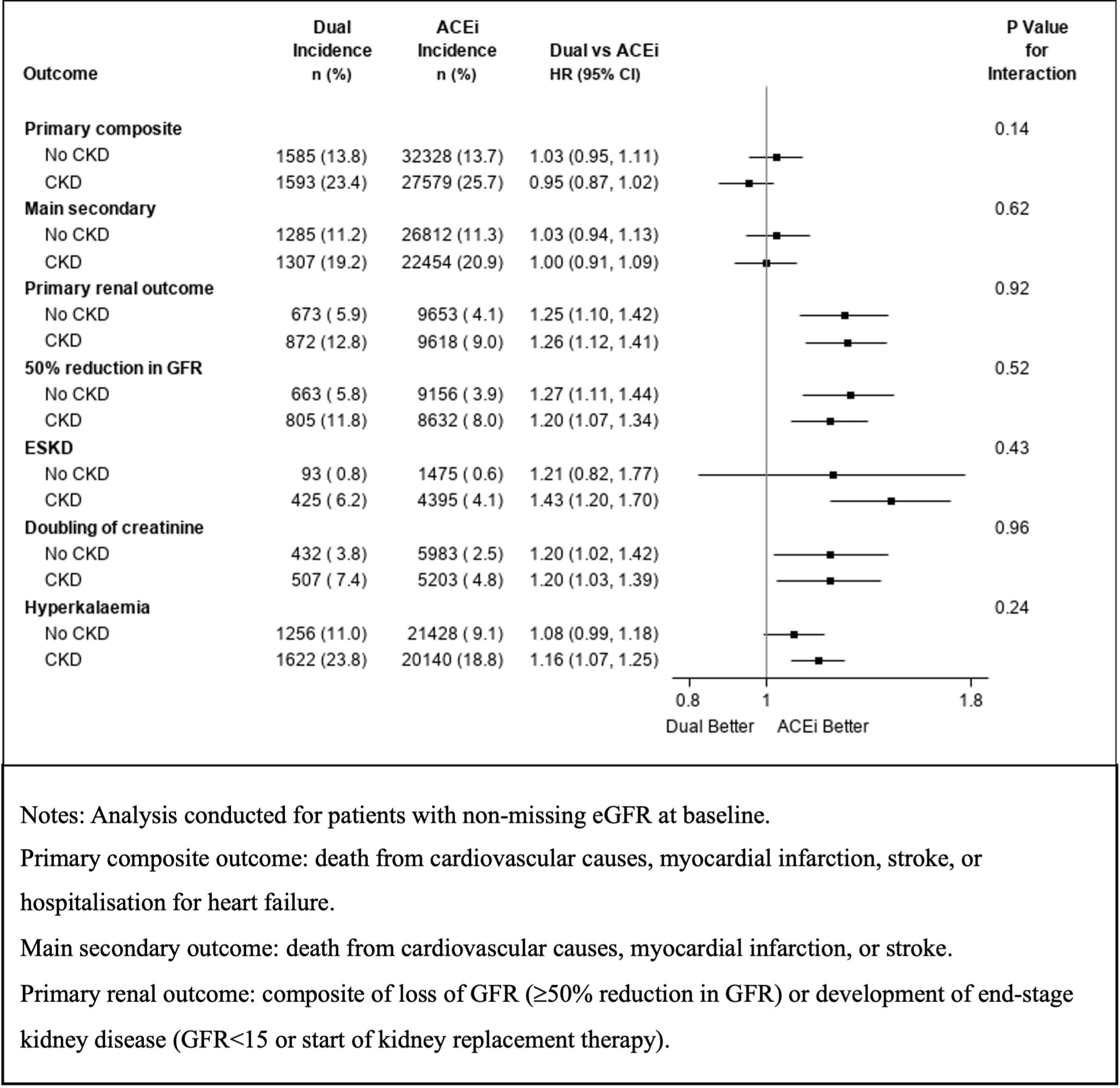

#### Sensitivity analyses

We observed that dual therapy patients included in alternate definition 1 had higher blood pressure at baseline and had more GP and hospital appointments within the 6 months prior to follow-up, compared to the primary definition used in the main analysis and alternate definition 2 (Supplementary Table S6). Using the dual therapy alternate definition 1 described in the Appendix and displayed in Supplementary Figure S2, which may introduce immortal-time bias and selection bias, the risk of the primary composite cardiovascular outcome was increased among dual therapy users compared to ACEi users, HR 1.15 (95% CI: 1.09, 1.22). An increased risk was also observed for dual therapy vs ACEi use for the main secondary outcome, which differed from the main analysis results using the primary definition and the ONTARGET trial results (Supplementary Figure S3). When using alternate definition 2 for dual therapy users, results for the primary composite cardiovascular outcome were similar to the main analysis and consistent with the ONTARGET trial findings, HR 0.97 (95% CI: 0.93, 1.02). Results were also similar for the main secondary outcome. However, the HR for all outcomes estimated a slightly lower HR effect size when compared to the primary definition used in the main analysis and ONTARGET results (Supplementary Figure S3).

After imputation of baseline blood pressure and creatinine and re-regeneration of propensity-score—weights, we estimated similar results for dual therapy vs ACEi for the primary composite outcome as in the main analysis and consistent with the ONTARGET trial results, HR 0.97 (95% CI: 0.92, 1.02).

28.9% of the patients included in analysis had albumin-creatinine ratio at baseline. Among those with non-missing values, we found no statistically significant interaction for renal outcomes and hyperkalaemia when stratifying by baseline proteinuria status. However, dual therapy use was associated with an increased risk of cardiovascular outcomes for patients with proteinuria (Supplementary Figure S4).

Dual therapy patients had a longer duration of exposure to antihypertensives prior to the start of follow up compared to ACEi patients. After adjusting for length of time since first exposure to any antihypertensive (ARB/ACEi/dual therapy) the HR was 0.96 (95% CI: 0.90, 1.00) for the primary composite outcome, similar to the main analysis.

## DISCUSSION

In this emulation of the dual therapy arm of the ONTARGET randomised trial, using a large routinely-collected healthcare dataset, we found no difference in treatment effects of dual therapy and ACEi use for the primary and secondary composite cardiovascular outcomes, consistent with the trial. We found dual therapy was associated with an increased risk of ESKD or ≥50% reduction in GFR when assessed separately and as a composite outcome, consistent with the findings of the renal outcomes studied in ONTARGET, despite slight differences in the outcome definitions. We also found results consistent with the trial for doubling of serum creatinine and observed similar conclusions for the outcome of hyperkalaemia, both of which were increased with dual therapy compared with ACEi treatment alone.

After successfully benchmarking results for the primary composite cardiovascular outcome and aided by greater power, including an increased number of people with CKD at baseline, we were able to extend findings and examine for treatment effect heterogeneity by baseline CKD status. We found no evidence of treatment effect heterogeneity by CKD for any of the outcomes.

We also examined the reliability of our primary operational definition of dual therapy use in routine care data by comparing results to those obtained in the ONTARGET trial. We started follow-up for dual therapy users at two additional timepoints and assessed the impact of different sources of bias from varying definitions of dual use on the results.

### Strengths and Limitations

After benchmarking findings against the ONTARGET trial, we observed that trial results for the cardiovascular and renal outcomes extended to patients with CKD at baseline who were underrepresented in ONTARGET.

Although prescribing of dual therapy is no longer recommended, uncertainty remains about the balance of risks and benefits in subgroups or as an individualisation strategy.^29^ Understanding the potential harms of therapy in routine care, where patient monitoring is substantially less rigorous than in a clinical trial, is therefore important. In part because guidance has recommended against dual RAS blockade since 2009, the number of dual users in routine care over this time period is low. Many of these would not have been detected in our analysis by requiring initial prescriptions on the same day to define dual use: the initial decision to start dual blockade may not have resulted in simultaneous prescriptions if patients were already taking one agent or if the therapeutic intent was to start two agents but to gradually uptitrate. Our pragmatic definition of users of dual blockade maximised power and avoided omitting some of the early follow-up time but could have resulted in survivor bias. Therefore, we robustly assessed the impact of our definition of dual users in sensitivity analyses. Our findings demonstrate that the results are sensitive to how dual therapy use is defined in routine care data. Fewer events in the dual therapy arm and a conservative effect of dual users were expected for the definition where survivor bias was increased, however for this analysis we obtained results consistent with the trial indicating that the impact of this bias was likely to be minimal. On the other hand, we observed when using a definition which is likely to introduce immortal time bias and selection bias (alternate definition 1), results were showed an increased risk associated with dual therapy use for all outcomes not consistent with the ONTARGET trial results. This could be due to confounding by indication with patients having an event before they receive a subsequent prescription for the 2^nd^ agent.

Furthermore, the mean number of days between the start of follow up in alternate definition 1 compared to start of follow up in the primary definition used in the main analysis, (i.e., between the 1^st^ prescription for the 2^nd^ agent and the 2^nd^ prescription for the 1^st^ agent) was 185 days. Due to dual users in alternate definition 1 starting follow-up from an earlier point compared to the main definition and alternate definition 2, these patients were exposed for a shorter period. Therefore, patients in this analysis who may have had an outcome such as stroke or heart failure would have subsequently been excluded from analysis using the definition used in the main analysis and alternate definition 2, as exclusion criteria were reassessed at start of follow up. This indicates that the issues of bias and confounding, which will be specific to individual therapeutic areas, need to be carefully considered by research teams studying this question in the future.

We identified that patients being prescribed dual therapy were more ethnically diverse, with higher blood pressure, higher baseline creatinine and a higher proportion of people with high-risk diabetes compared with those prescribed an ACEi alone. Despite this, by using an operational definition, we were able to replicate the primary and secondary outcomes of ONTARGET in our emulation, providing confidence that the RCT results are generalisable to this wider routine-care population. We also demonstrated an increased risk of renal outcomes among dual therapy compared with ACEi users, adding evidence to the generalizability of the ONTARGET trial findings. This is consistent with previous study findings and supports the recommendation against the use of dual therapy.^1, 3^ However, a greater proportion of patients who met trial criteria receiving dual therapy were being treated for proteinuria in the context of diabetes (19.8%), compared with patients receiving ACEi alone (11.9%). These results do not support the hypothesis that dual therapy may provide renal protection in those with CKD at baseline; however, we recognize that they may also be due to confounding by indication. Evidence against this is our finding that, for renal outcomes, results were consistent when stratified by baseline proteinuria status. Despite this being the first study to our knowledge to assess heterogeneity of dual therapy use by proteinuria status, due to the limited number of patients with album-creatinine ratio measures recorded at baseline, these results should be interpreted with caution. Observed baseline differences in indication may have also contributed to the higher risk of the composite renal outcome observed in the dual therapy arm, since dual blockade was indicated for treatment of progressive proteinuric CKD for much of the time period of this study.^30–32^ Our results from benchmarking against the trial were consistent with ONTARGET findings where confounding by indication was not present due to randomisation.

There was a substantial amount of missing data for blood pressure and creatinine which could have led to bias. However, after multiple imputation under the assumption these variables were missing at random given the other covariates, the outcomes and the exposure, results were consistent with the results of the main analysis.

### Comparison to other studies

Our results suggest that dual ACEi/ARB therapy was associated with an increased risk of renal outcomes compared to ACEi alone in agreement with a smaller observational study by Caravaca-Fontán et al., which found dual therapy was associated with a faster decline of renal function in patients with CKD.^33^ Fralick et al.^34^ used US insurance claims data to replicate ONTARGET results for the single therapy comparison and led to results closely comparable to the trial for 9330 patients but omitted the dual therapy analysis from their replication. Methodologically, although an increasing number of studies have used trial emulation methods across various therapeutic areas, few have looked at how to define dual therapy use in routine data. A study exploring treatment for breast cancer using trial replication methods by Merola et al., included dual users as patients who received prescriptions for both drugs on the same day.^35^ To our knowledge this is the first study exploring the feasibility of trial replication methods applied to dual therapy treatment arms for cardiovascular disease. In this therapeutic area and the management of hypertension, selection bias may be introduced when restricting the cohort to users with simultaneous prescriptions as many patients may be initiated on dual therapy as add-ons to an existing medication. Our work suggests that using a practical proxy definition of a dual user when the sample of actual dual therapy users is small may be sufficient to reproduce trial findings. Further research using more complex trial emulation methods, such as the clone-censor-weight method,^36^ would represent an interesting area for future research.

### Conclusion

In this emulation of the dual therapy arm of the ONTARGET randomised trial using routinely-collected healthcare data, we confirmed similar effectiveness of dual therapy compared to ACEi alone at reducing the risk of a composite of cardiovascular death, MI, stroke or hospital admission for congestive heart failure. Also consistent with the trial, we observed increased risk for the outcome of doubling of serum creatinine, and for a renal composite outcome of ≥50% reduction in GFR or ESKD, and for hyperkalaemia, among dual therapy users compared with users of ACEi alone. Cardiovascular results extended to patients with CKD at baseline who were underrepresented in the trial, with no evidence of heterogeneity by CKD status.

This study demonstrates that a dual therapy arm can be emulated using observational data, but highlights the importance of considering potential sources of bias that may be introduced depending on how dual therapy is defined. These considerations will be specific to each therapeutic area and research question.

## DISCLOSURE

PB was supported by Barts Charity (MGU0504) at the time of the review and was funded by a GSK PhD studentship at the time of analysis and writing. AYSW was supported by the British Health Foundation fellowship. AS is employed by LSHTM on a fellowship sponsored by GSK. EP was an employee of and holds stock in Compass Pathways at the time of the review. CC has received consultation, advisory board membership or research funding from the Ontario Ministry of Health, Sanofi, Pfizer, Leo Pharma, Astellas,

Janssen, Amgen, Boehringer-Ingelheim and Baxter. In 2018 CC co-chaired a KDIGO potassium controversies conference sponsored at arm’s length by Fresenius Medical Care, AstraZeneca, Vifor Fresenius Medical Care, Relypsa, Bayer HealthCare and Boehringer Ingelheim. CC co-chairs the cloth mask knowledge exchange, a stakeholder group that includes cloth mask manufacturers and fabric distributors. JFEM reports honoraria from AstraZeneca, Bayer, Boehringer, Novo Nordisk, UpToDate

Inc., Idorisia, Labchem, Parexel, Roche, Sanofi. MC was an employee of GSK at the time of the study. All other authors have no conflicts.

## DATA SHARING

This study is based in part on data from the Clinical Practice Research Datalink obtained under licence from the UK Medicines and Healthcare products Regulatory Agency (protocol no. 20_012). The data is provided by patients and collected by the NHS as part of their care and support. Ethical approval has been granted by the London School of Hygiene & Tropical Medicine Ethics Committee (Ref: 22658). The study has been approved by the Independent Scientific Advisory Committee of the UK Medicines and Healthcare Products Regulatory Agency (protocol no. 20_012). Access to the individual patient data from the ONTARGET trial was obtained by the trial investigators and complies with institutional review board approved informed consent forms provided by the individuals from whom the data were collected but is not publicly available. Code lists used to develop the study population and define outcomes are available for download: https://doiorg/1017037/DATA00002112.

## ACKNOWLEDGMENTS

This work was supported by the funding from a GlaxoSmithKline PhD studentship held by PB as part of an ongoing collaboration between GSK and the London School of Hygiene and Tropical Medicine. This research was funded in whole, or in part, by the Wellcome Trust [Senior Research Fellowship 224485/Z/21/Z]. For the purpose of open access, the author has applied a CC BY public copyright licence to any Author Accepted Manuscript version arising from this submission. The funders of the study had no role in the study design, data collection, data analysis, data interpretation, or writing of the report. All authors had full access to all the data and take final responsibility for the decision to submit for publication. We are grateful to the ONTARGET trial sponsors for providing access to the trial data to facilitate this study and to the patients who contribute their data to the Clinical Practice Research Datalink. All authors contributed to the conceptualisation, study question and design. PB, LT, KW, EW, AW, AS, CL, CC and EP contributed to the statistical analysis decisions of this study. PB completed formal analysis of this study and wrote the original draft of this manuscript under the supervision of LT, KW and MC. PB, LT, KW, EW, AW, AS, CL, EP had access to the data and data reported in this manuscript was verified by LT and KW. All authors contributed to the writing, review and editing of the submitted manuscript.

