## Appendix for "Cardiorenal effects of dual blockade with Angiotensin-converting enzyme inhibitors and Angiotensin receptor blockers in people with CKD: analysis of routinely collected data with emulation of a reference trial (ONTARGET)"

We assessed bias introduced in our primary operational definition of dual users through two alternate dual therapy definitions described below:

- **Alternate definition 1: Starting follow-up at the date of the 1st prescription for the 2nd agent**

To explore the impact of the bias from excluding those who may die early or have early adverse events, described above in the operational definition, we started follow-up at the date of 1st prescription for the 2nd agent (Supplementary Figure S2). The trial eligibility criteria were reassessed at the new follow-up start time. This reduced the element of survivor bias. However, because only those patients who met the operational definition were included, this introduces a period during which a fatal event could not occur between the new start of follow-up and when the operational definition was met, which leads to immortal time bias. Additionally, selection bias may be present as patients included in this definition are likely to have had less exposure to medication.

- **Alternate definition 2: Starting follow-up at the date of the 2nd prescription for the 2nd agent**

Additionally, to assess the trade-off between survivor and immortal time bias we started follow-up from the date of 2nd prescription for the 2nd agent (Supplementary Figure S2), reassessing trial eligibility criteria at this time point. Immortal time bias is still present in this definition, but reduced compared with alternate definition 1 and the duration of survivor bias was also reduced compared with the primary operational definition. In comparison to the first alternate definition, starting follow up at this point would mean patients may be likely to have been receiving medication for a longer period, similar to the operational definition.

Eligibility criteria were the same as the ONTARGET trial and were reassessed at the start of start of follow-up for the two alternate dual therapy definitions, generating trial-eligible periods.
