## Supplementary figures and images for "Cardiorenal effects of dual blockade with Angiotensin-converting enzyme inhibitors and Angiotensin receptor blockers in people with CKD: analysis of routinely collected data with emulation of a reference trial (ONTARGET)"

### Supplementary Figure S1

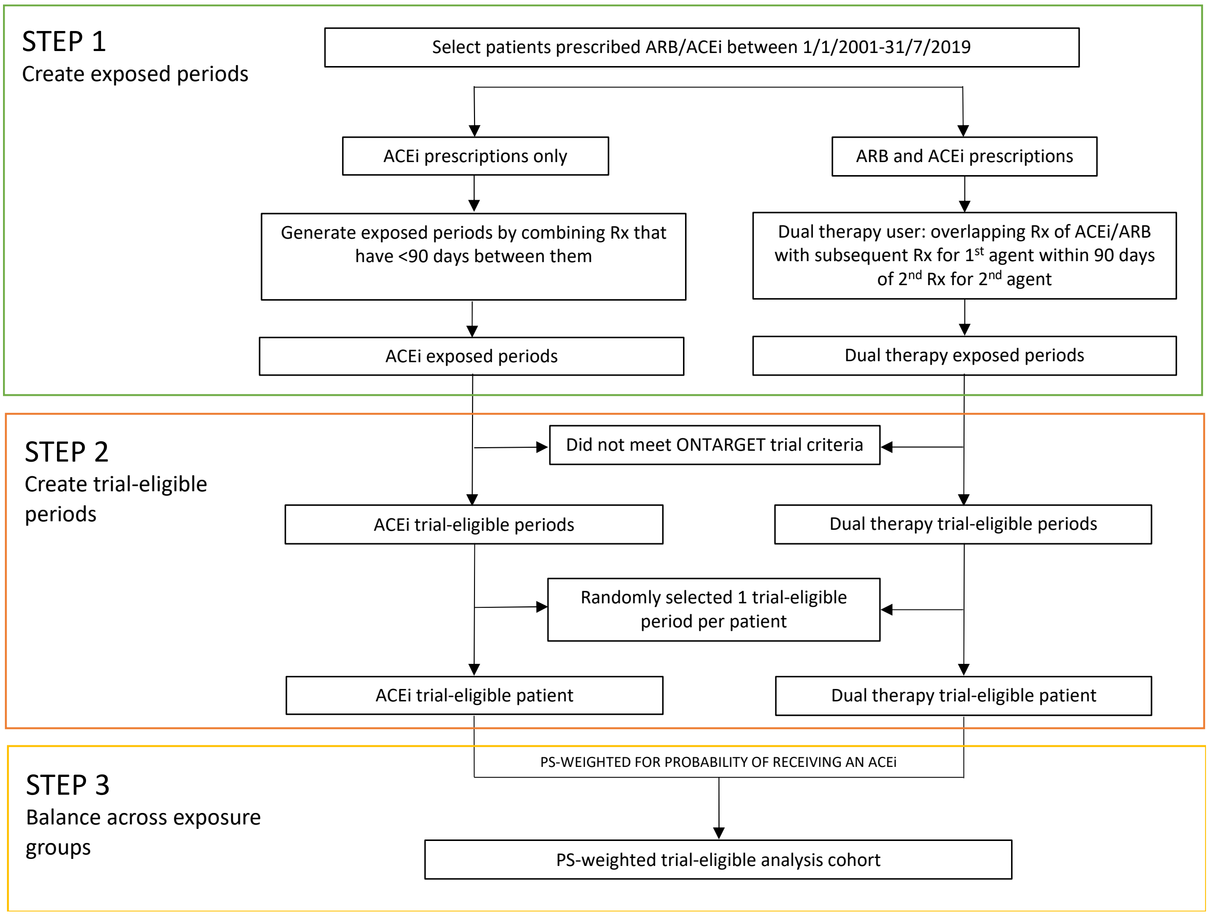


**Supplementary Figure S1.** Steps to define analysis cohort.

Rx=prescription
