## Supplementary Figure S2 for "Cardiorenal effects of dual blockade with Angiotensin-converting enzyme inhibitors and Angiotensin receptor blockers in people with CKD: analysis of routinely collected data with emulation of a reference trial (ONTARGET)"

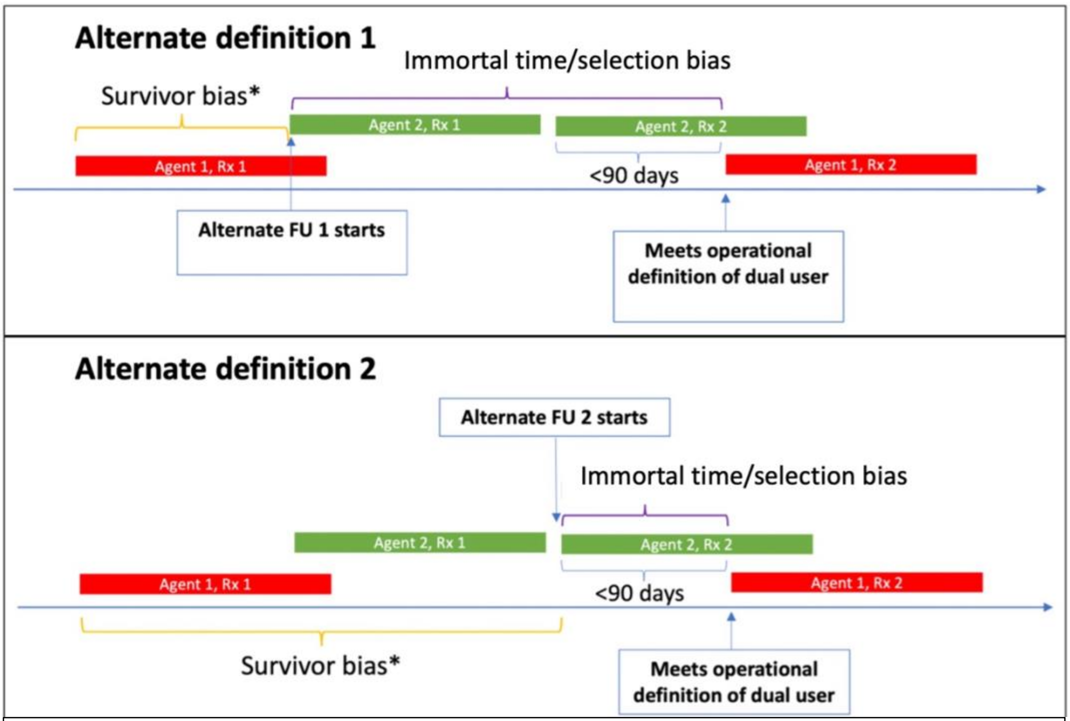


Notes: Dual users are patients with overlapping prescriptions (<90 days between prescriptions) who have subsequent prescription for 1st agent within 90 days of 2nd prescription for 2nd agent. Alternate definition 1: follow-up started from date of 1st prescription for 2nd agent; Alternate definition 2: follow-up started from date of 2nd prescription for 1st agent. Agent=medication; Rx=prescription; FU=follow-up for dual user. Survivor bias* represents bias introduced from operational definition excluded patients who may have event or die early.

**Supplementary Figure S2.** Illustration of alternate definitions of a dual user and potential biases
