## Supplementary Figure S3 for "Cardiorenal effects of dual blockade with Angiotensin-converting enzyme inhibitors and Angiotensin receptor blockers in people with CKD: analysis of routinely collected data with emulation of a reference trial (ONTARGET)"

| Outcome | Dual Incidence n (%) | ACEi Incidence n (%) | Dual vs ACEi HR (95% CI) |
|---|---|---|---|
| **Primary composite** | | | |
| ONTARGET | 1386 (16.3) | 1412 (16.5) | 0.99 (0.92, 1.07) |
| Primary definition | 3478 (17.5) | 69258 (17.6) | 0.98 (0.93, 1.03) |
| Alternate definition 1 | 3065 (23.4) | 90014 (19.6) | 1.15 (1.09, 1.22) |
| Alternate definition 2 | 3794 (18.5) | 76591 (18.7) | 0.97 (0.93, 1.02) |
| **Main secondary** | | | |
| ONTARGET | 1200 (14.1) | 1210 (14.1) | 1.00 (0.93, 1.09) |
| Primary definition | 2853 (14.3) | 57220 (14.6) | 1.01 (0.95, 1.07) |
| Alternate definition 1 | 2377 (18.2) | 73607 (16.0) | 1.08 (1.01, 1.16) |
| Alternate definition 2 | 3106 (15.1) | 62999 (15.4) | 0.99 (0.95, 1.04) |
| **Primary renal outcome** | | | |
| ONTARGET | 212 ( 2.5) | 174 ( 2.0) | 1.24 (1.01, 1.51) |
| Primary definition | 1545 ( 8.5) | 19271 ( 5.6) | 1.25 (1.15, 1.36) |
| Alternate definition 1 | 1424 (12.2) | 23832 ( 5.9) | 1.39 (1.27, 1.52) |
| Alternate definition 2 | 1708 ( 9.2) | 21274 ( 5.9) | 1.11 (1.04, 1.19) |
| **50% reduction in GFR** | | | |
| ONTARGET | - | - | - |
| Primary definition | 1468 ( 8.0) | 17788 ( 5.2) | 1.23 (1.13, 1.34) |
| Alternate definition 1 | 1259 (10.7) | 21917 ( 5.5) | 1.33 (1.21, 1.47) |
| Alternate definition 2 | 1550 ( 8.4) | 19558 ( 5.4) | 1.10 (1.02, 1.18) |
| **ESKD** | | | |
| ONTARGET | - | - | - |
| Primary definition | 578 ( 2.9) | 6733 ( 1.7) | 1.38 (1.19, 1.60) |
| Alternate definition 1 | 710 ( 5.4) | 8377 ( 1.8) | 1.67 (1.47, 1.88) |
| Alternate definition 2 | 709 ( 3.5) | 7438 ( 1.8) | 1.23 (1.11, 1.37) |
| **Doubling of creatinine** | | | |
| ONTARGET | 166 ( 2.0) | 140 ( 1.6) | 1.20 (0.96, 1.50) |
| Primary definition | 939 ( 5.1) | 11186 ( 3.3) | 1.20 (1.07, 1.34) |
| Alternate definition 1 | 825 ( 7.0) | 13907 ( 3.5) | 1.41 (1.24, 1.59) |
| Alternate definition 2 | 1013 ( 5.5) | 12337 ( 3.4) | 1.06 (0.98, 1.16) |
| **Hyperkalaemia** | | | |
| ONTARGET | - | - | 1.58 (1.26, 1.99) |
| Primary definition | 2922 (14.7) | 42020 (10.7) | 1.15 (1.09, 1.22) |
| Alternate definition 1 | 2197 (16.8) | 49442 (10.8) | 1.24 (1.15, 1.32) |
| Alternate definition 2 | 3054 (14.9) | 45089 (11.0) | 1.06 (1.01, 1.11) |

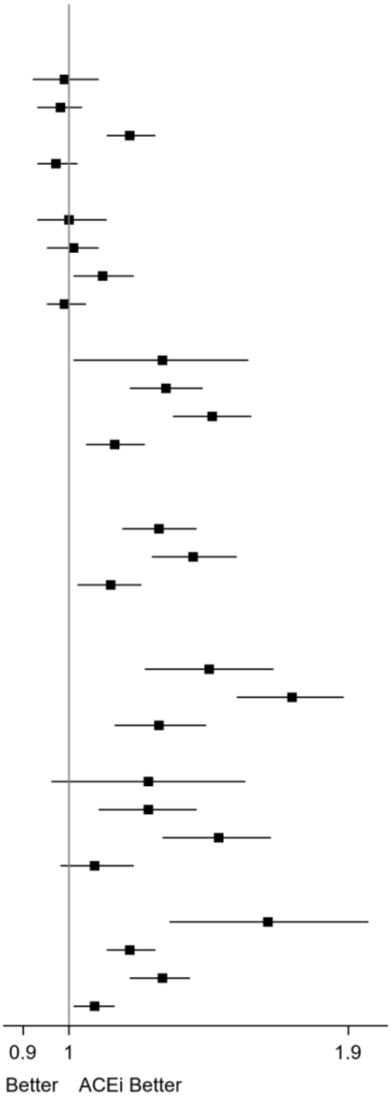


Notes: Dual therapy users defined as patients with overlapping prescriptions (<90 days between prescriptions) who have subsequent prescription for 1st agent within 90 days of 2nd prescription for 2nd agent. Primary definition (main analysis): follow-up started from date definition met i.e., date of 2nd prescription for 1st agent. Alternate definition 1: follow-up started from date of 1st prescription for 2nd agent. Alternate definition 2: follow-up started from date of 2nd prescription for 2nd agent. Renal outcomes (except ESKD) studied among patients with non-missing creatinine at baseline.

Primary composite outcome: death from cardiovascular causes, myocardial infarction, stroke, or hospitalisation for heart failure.

Main secondary outcome: death from cardiovascular causes, myocardial infarction, or stroke.

Primary renal outcome: composite of loss of GFR (≥50% reduction in GFR) or development of end-stage kidney disease (GFR<15 or start of kidney replacement therapy).

ESKD: GFR<15 or start of kidney replacement therapy.

ONTARGET studied hyperkalaemia at 6 weeks therefore not directly comparable to our results which studied hyperkalaemia during total follow up.

**Supplementary Figure S3.** Forest plot of results for primary operational definition and alternate definitions of dual users
