## Supplementary Figure S4 for "Cardiorenal effects of dual blockade with Angiotensin-converting enzyme inhibitors and Angiotensin receptor blockers in people with CKD: analysis of routinely collected data with emulation of a reference trial (ONTARGET)"

| Outcome | Dual Incidence n (%) | ACEi Incidence n (%) | Dual vs ACEi HR (95% CI) | P Value for Interaction |
|---|---|---|---|---|
| **Primary composite** | | | | 0.01 |
| No proteinuria | 504 (14.4) | 10194 (15.1) | 0.87 (0.76, 1.00) | |
| Proteinuria | 779 (23.6) | 9948 (22.3) | 1.13 (1.00, 1.28) | |
| **Main secondary** | | | | 0.05 |
| No proteinuria | 421 (12.0) | 8330 (12.3) | 0.91 (0.78, 1.05) | |
| Proteinuria | 597 (18.1) | 7976 (17.9) | 1.12 (0.97, 1.29) | |
| **Primary renal outcome** | | | | 0.93 |
| No proteinuria | 242 ( 7.0) | 3190 ( 4.7) | 1.28 (1.03, 1.60) | |
| Proteinuria | 488 (15.0) | 4673 (10.6) | 1.27 (1.08, 1.49) | |
| **50% reduction in GFR** | | | | 0.75 |
| No proteinuria | 233 ( 6.7) | 2973 ( 4.4) | 1.30 (1.04, 1.62) | |
| Proteinuria | 464 (14.3) | 4287 ( 9.7) | 1.24 (1.05, 1.46) | |
| **ESKD** | | | | 0.37 |
| No proteinuria | 56 ( 1.6) | 809 ( 1.2) | 1.13 (0.70, 1.83) | |
| Proteinuria | 186 ( 5.6) | 1642 ( 3.7) | 1.46 (1.10, 1.92) | |
| **Doubling of creatinine** | | | | 0.49 |
| No proteinuria | 135 ( 3.9) | 1798 ( 2.7) | 1.19 (0.89, 1.60) | |
| Proteinuria | 332 (10.2) | 2866 ( 6.5) | 1.35 (1.10, 1.65) | |
| **Hyperkalaemia** | | | | 0.20 |
| No proteinuria | 575 (16.4) | 8983 (13.3) | 1.07 (0.95, 1.22) | |
| Proteinuria | 833 (25.2) | 9058 (20.3) | 1.20 (1.07, 1.35) | |

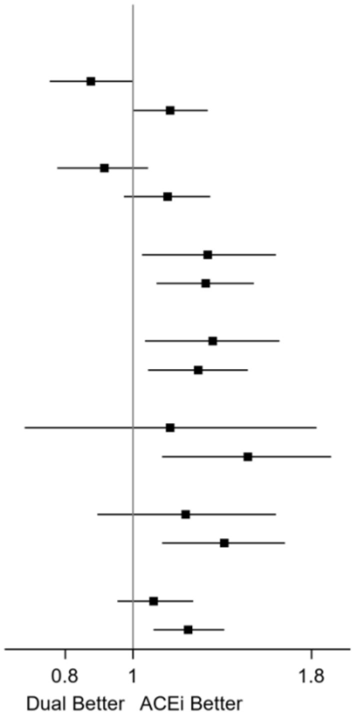


Notes: Analysis conducted for patients with non-missing albumin-creatinine ratio at baseline. Renal outcomes (except ESKD) studied among patients with non-missing creatinine at baseline.

Primary composite outcome: death from cardiovascular causes, myocardial infarction, stroke, or hospitalisation for heart failure.

Main secondary outcome: death from cardiovascular causes, myocardial infarction, or stroke.

Primary renal outcome: composite of loss of GFR (≥50% reduction in GFR) or development of end-stage kidney disease (GFR<15 or start of kidney replacement therapy).

ESKD: GFR<15 or start of kidney replacement therapy.

**Supplementary Figure S4.** Forest plot of test for treatment effect heterogeneity by proteinuria status at baseline.
