## Supplementary Table S1 for "Cardiorenal effects of dual blockade with Angiotensin-converting enzyme inhibitors and Angiotensin receptor blockers in people with CKD: analysis of routinely collected data with emulation of a reference trial (ONTARGET)"

**Supplementary Table S1**. Table of key design aspects of the ONTARGET trial dual analysis and emulation in CPRD Aurum

| **Protocol component** | **ONTARGET** | **Trial emulation protocol** | **Implementation in CPRD Aurum** |
|---|---|---|---|
| Eligibility criteria | Patients aged ≥55 years with coronary artery, peripheral artery or cerebrovascular disease or high-risk diabetes with end organ damage recruited up to 2004. No restriction on previous ACEi/ARB use except must be able to discontinue use. | Patients with a prescription for an ACEi or ARB between 01 January 2001 to 31 July 2019, eligible for HES linkage, aged ≥55 years with coronary artery, peripheral vascular, or cerebrovascular disease or high-risk diabetes. | As in protocol. |
| Treatment strategies | Patients entered 3-week single blind run-in period to check compliance then randomised to one of three trial arms: ramipril 10 mg + telmisartan placebo, telmisartan 80 mg + ramipril placebo or ramipril 10 mg + telmisartan 80 mg. | Continuous courses of therapy with treatment gaps of <90 days. Dual therapy users defined as patients with overlapping prescriptions who received a subsequent prescription for the 1st agent after the 2nd prescription for the 2nd agent. | As in protocol with condition added for dual therapy definition that subsequent prescription for 1st agent must occur within 90 days of 2nd prescription for 2nd agent to avoid capturing treatment switchers, i.e., patients with overlapping prescriptions who received a subsequent prescription for the 1st agent with 90 days of the 2nd prescription for the 2nd agent. |
| Assignment procedures | Randomly assigned and received placebo for other drug so unaware which arm assigned to | Based on prescriptions received. Patient could contribute to both exposure groups at different timepoints | As in protocol. |
| Follow-up period | Follow-up started at randomisation and ended at primary event, death, loss to follow-up or end of study. Close out was planned in July 2007. | Follow-up started at start of trial-eligible period where exposure period met trial inclusion/exclusion criteria. For dual users follow-up started at date of 1st prescription for 2nd agent. | As in protocol but for dual users follow-up started at date meets criteria for dual user, i.e., the date of the 2nd prescription for the 1st agent, conditional on meeting the trial criteria. |

**Supplementary Table S1**. Table of key design aspects of the ONTARGET trial dual analysis and emulation in CPRD Aurum

| **Protocol component** | **ONTARGET** | **Trial emulation protocol** | **Implementation in CPRD Aurum** |
|---|---|---|---|
| | | Ended at the earliest of: outcome of interest, death, transferred out of practice date, or last data collection from the general practice. If these dates did not occur the patient was censored after 5.5 years of follow-up | Impact of this choice of start of follow-up assessed in sensitivity analysis. |
| Outcome | Primary composite of: cardiovascular death, MI, stroke, hospitalisation for heart failure | As in ONTARGET, defined using ICD10, Read codes and death registries from ONS. | As in protocol. |
| Analysis plan | Primary analysis under time-to-event counting first occurrence of any component of the composite outcome using Cox proportional hazards model. Intention-to-treat as main analysis | Match to trial to obtain trial-analogous cohort then will match trial-eligible exposure groups. Cox proportional hazards model used for primary analysis. | Analysis conducted on one randomly selected trial eligible period per patient. Balance of covariates obtained by propensity score weighting for probability of receiving an ACEi. Weighting as opposed to matching to increase sample size and methods trialled and led to comparable results in replication of single therapy analysis. Cox proportional hazards model used for primary analysis. |
