## Supplementary Table S2 for "Cardiorenal effects of dual blockade with Angiotensin-converting enzyme inhibitors and Angiotensin receptor blockers in people with CKD: analysis of routinely collected data with emulation of a reference trial (ONTARGET)"

**Supplementary Table S2.** Table of trial diagnoses (inclusion criteria) and interpretation in CPRD.

| **ONTARGET/TRANSCEND** | **CPRD Aurum (HES + ONS Linked)**<br>**READ or ICD 10 code for:** |
|---|---|
| Aged ≥55 years | Aged ≥55 years prior to prescription of drug |
| Coronary artery disease | |
| Previous myocardial infarction (>2 days post uncomplicated MI) | MI at least 2 days prior to prescription of drug |
| Stable angina or unstable angina >30 days before informed consent and with documented evidence of multivessel coronary artery disease | Angina/stable angina/unstable angina at least 30 days before prescription of drug and previous coronary artery disease diagnosis |
| Multi-vessel PTCA >30 days before informed consent | Read, ICD-10 or OPCS code for coronary angioplasty at least 30 days before prescription of drug |
| Multi-vessel CABG surgery >4 years before informed consent, or with recurrent angina following surgery | Read, ICD-10 or OPCS code for CABG at least 4 years before prescription of drug or with angina after CABG |
| Peripheral artery disease | |
| Previous limb bypass surgery or angioplasty | Read, ICD-10 or OPCS code for limb bypass surgery or angioplasty |
| Previous limb or foot amputation | Read, ICD-10 or OPCS code for limb/foot amputation |
| Intermittent claudication, with ankle:arm BP ratio <=0.80 on at least 1 side | Intermittent claudication |
| Significant peripheral artery stenosis (>50%) documented by angiography or non-invasive test | Not applicable |
| Cerebrovascular disease | |
| Previous stroke | Stroke before prescription of drug |
| Transient ischemic attacks >7 days and <1 year before informed consent | Transient ischemic attacks before prescription of drug |
| High-risk diabetes with evidence of end-organ damage | |
| High-risk diabetes | Specific codes for diabetes with retinopathy, neuropathy, chronic kidney disease or proteinuria |

| | |
|---|---|
| | before prescription of drug or diabetes defined by diabetes codes or diabetes therapy with CKD defined as eGFR<60 or proteinuria defined as ACR>3 |

Notes: Where dates are used as criteria dates from both CPRD and HES will be used, but if available HES will be preferred.
