## Supplementary Table S3 for "Cardiorenal effects of dual blockade with Angiotensin-converting enzyme inhibitors and Angiotensin receptor blockers in people with CKD: analysis of routinely collected data with emulation of a reference trial (ONTARGET)"

**Supplementary Table S3.** Table of trial exclusion criteria and interpretation in CPRD.

| **ONTARGET/TRANSCEND exclusion criteria** | **CPRD Aurum (HES + ONS Linked)**<br>**READ or ICD 10 code (prior to eligible for inclusion date, unless otherwise specified) for:** |
|---|---|
| Inability to discontinue ACEi or ARB | Not applicable |
| Known hypersensitivity or intolerance to ACEi or ARB | Not applicable |
| Symptomatic congestive heart failure | Heart failure or left ventricular dysfunction |
| Hemodynamically significant primary valvular or outflow tract obstruction | Aortic or pulmonary stenosis or previous valve replacement |
| Constrictive pericarditis | Constrictive pericarditis |
| Complex congenital heart disease | Congenital heart disease |
| Syncopal episodes of unknown etiology <3 months before informed consent | Not applicable |
| Planned cardiac surgery or PTCA <3 months of informed consent | Not applicable |
| Uncontrolled hypertension on treatment (e.g. BP >160/100 mm Hg) | Last recorded BP >160/100 mmHg for patients on treatment with other antihypertensives prior to ACEi/ARB initiation |
| Heart transplant recipient | Read, ICD-10 or OPCS code for heart transplant recipient |
| Stroke due to subarachnoid haemorrhage | Previous cerebral haemorrhage |
| Significant renal artery disease | Codes for renal artery stenosis or renal artery atherosclerosis; or serum creatinine concentration above 265μmol/L |
| Hepatic dysfunction | Cirrhosis or other documented liver disease |
| Uncorrected volume or sodium depletion | Not applicable |
| Primary hyperaldosteronism | Primary hyperaldosteronism/ Conn's syndrome |
| Hereditary fructose intolerance | Hereditary fructose intolerance |

| | |
|---|---|
| Other major noncardiac illness expected to reduce life expectancy or interfere with study participation | Recorded solid organ or metastatic malignancy within the last 5 years, drug, alcohol dependence or mental illness. |
| Simultaneously taking another experimental drug | Not applicable |
| Significant disability precluding regular follow-up visits | Not applicable |
| Unable or unwilling to provide written informed consent | Not applicable |
| Elevated potassium above 5.5mmol/L | Elevated potassium above 5.5mmol/L |
| Hypotension | SBP <90 mm Hg |

Notes: Where dates are used as criteria dates from both CPRD and HES will be used, but if available HES will be preferred. Not applicable used when anticipated there will be extensive missing data or risk of misclassification.
