## Supplementary Table S4 for "Cardiorenal effects of dual blockade with Angiotensin-converting enzyme inhibitors and Angiotensin receptor blockers in people with CKD: analysis of routinely collected data with emulation of a reference trial (ONTARGET)"

**Supplementary Table S4.** List of variables considered and included in propensity-score model

| **Potential confounders** | **Selected into propensity-score model** | **Reason for omitting** |
|---|---|---|
| Stroke/TIA | ✓ | |
| Peripheral artery disease | ✓ | |
| Coronary artery disease | ✓ | |
| Diabetes | ✓ | |
| High-risk diabetes | ✓ | |
| Age (years) | ✓ | |
| Sex | ✓ | |
| Ethnicity | ✓ | |
| BMI | ✓ | 7.6% missing |
| SBP | ✓ | 22.5% missing |
| DBP | ✓ | 22.5% missing |
| Creatinine | ✓ | 19.5% missing |
| Index of Multiple Deprivation (IMD) | ✓ | 0.1% missing |
| Smoke status | ✓ | 2.7% missing |
| Alcohol use | | 16.9% missing |
| Statin use | ✓ | |
| Nitrate use | ✓ | |
| Diabetic treatment use | ✓ | |
| Diuretic use | ✓ | |
| CCB use | ✓ | |
| Betablocker use | ✓ | |
| Aspirin use | ✓ | |
| Antiplatelet use | ✓ | |
| Digoxin use | | Insufficient number of events |
| Anticoagulant use | ✓ | |
| Alpha-blocker use | ✓ | |
| No. of hospital admissions within 6 months prior | ✓ | |
| No. of GP appointments within 6 months prior | ✓ | |
| Year of start of eligible period | ✓ | |
| Time since first eligible period (days) | ✓ | |

Notes: TIA: transient ischaemic attack; BMI: body-mass index; SBP: systolic blood pressure; DBP: diastolic blood pressure.

Variables are measured at start of trial-eligible period or before.

Peripheral artery disease includes limb bypass surgery or angioplasty, limb/foot amputation, or intermittent claudication.

Coronary artery disease includes previous MI, angina, coronary angioplasty, or CABG.

SBP and DBP are measured within 6 months prior to start of trial-eligible period.

Medication use is within 3 months prior to start of trial-eligible period.

SBP and DBP had 22.5% missing but this variable was included as believed to be an important confounder. Similarly, baseline creatinine had 19.5% missing but due to the known differences in prescribing based on baseline creatinine this variable was also included in the model.

Balance after weighting was assessed for all variables listed including those not included in the propensity-score model but could be assumed to be missing at random. If imbalance remained analysis was adjusted for these variables.
