## Supplementary Table S5 for "Cardiorenal effects of dual blockade with Angiotensin-converting enzyme inhibitors and Angiotensin receptor blockers in people with CKD: analysis of routinely collected data with emulation of a reference trial (ONTARGET)"

**Supplementary Table S5.** Baseline characteristics and standardised differences of trial-eligible patients after applying trial criteria included in propensity-score—weighted analysis before and after weighting using operational definition of dual user

| **Characteristic** | **Before weighting** | | **After weighting** | | |
|---|---|---|---|---|---|
| | **Dual therapy** N=19 931 | **ACEi** N=392 475 | **Dual therapy** N=399 678 | **ACEi** N=412 551 | **SMD** |
| **Age (year) – mean (SD)** | 70.4 (8.8) | 70.5 (9.4) | 70.9 (9.4) | 70.5 (9.3) | 0.042 |
| **Systolic BP (mmHg) – mean (SD)** | 147.7 (20.0) | 144.3 (19.8) | 144.2 (19.5) | 144.5 (19.8) | 0.015 |
| **Diastolic BP (mmHg) – mean (SD)** | 79.7 (10.8) | 79.4 (10.9) | 79.4 (10.8) | 79.4 (10.9) | 0.002 |
| **Body-mass index – mean (SD)** | 29.7 (5.9) | 28.9 (5.8) | 29.0 (5.8) | 29.0 (5.8) | 0.001 |
| **Creatinine (µmol/l) – mean (SD)** | 96.4 (28.9) | 92.8 (26.8) | 92.3 (27.1) | 93.0 (27.0) | 0.024 |
| **Potassium (mmol/l) – mean (SD)** | 4.5 (0.5) | 4.4 (0.5) | 4.4 (0.5) | 4.4 (0.5) | 0.056 |
| **Female sex – no. (%)** | 10388 (52.1) | 186817 (47.6) | 203732 (51.0) | 197211 (47.8) | 0.063 |
| **Ethnic group – no. (%)** | | | | | |
| Black | 851 (4.3) | 10888 (2.8) | 11610 (2.9) | 11795 (2.9) | 0.003 |
| South Asian | 1565 (7.9) | 19152 (4.9) | 22117 (5.5) | 20603 (5.0) | 0.024 |
| White | 17186 (86.2) | 357202 (91.0) | 361266 (90.4) | 374531 (90.8) | 0.014 |
| Other | 329 (1.7) | 5233 (1.3) | 4685 (1.2) | 5622 (1.4) | 0.017 |
| **Clinical history – no. (%)** | | | | | |
| CAD[a] | 12296 (61.7) | 275353 (70.2) | 279174 (69.9) | 287704 (69.7) | 0.002 |
| Cerebrovascular disease[b] | 2149 (10.8) | 41967 (10.7) | 42336 (10.6) | 44125 (10.7) | 0.004 |
| PAD[c] | 1930 (9.7) | 36542 (9.3) | 36442 (9.1) | 38478 (9.3) | 0.007 |
| Diabetes | 13462 (67.5) | 237841 (60.6) | 241531 (60.4) | 251417 (60.9) | 0.010 |
| High-risk diabetes[d] | 12574 (63.1) | 189023 (48.2) | 192681 (48.2) | 201703 (48.9) | 0.014 |
| **Smoking status – no. (%)** | | | | | |
| Non-smoker | 6040 (30.3) | 106185 (27.1) | 111968 (28.0) | 112254 (27.2) | 0.018 |
| Current smoker | 4860 (24.4) | 105931 (27.0) | 106008 (26.5) | 110837 (26.9) | 0.008 |
| Past smoker | 9031 (45.3) | 180359 (46.0) | 181702 (45.5) | 189461 (45.9) | 0.009 |
| **Alcohol status – no. (%)** | | | | | |
| Non-drinker | 3711 (18.6) | 61706 (15.7) | 67423 (16.9) | 65393 (15.9) | 0.028 |
| Current drinker | 11578 (58.1) | 245276 (62.5) | 236509 (59.2) | 257249 (62.4) | 0.065 |
| Past drinker | 2647 (13.3) | 49349 (12.6) | 50986 (12.8) | 51983 (12.6) | 0.005 |
| Missing | 1995 (10.0) | 36144 (9.2) | 44761 (11.2) | 37926 (9.2) | 0.066 |
| **Medication[e] – no. (%)** | | | | | |
| Alpha-blocker | 2817 (14.1) | 35819 (9.1) | 37992 (9.5) | 38682 (9.4) | 0.004 |

**Supplementary Table S5.** Baseline characteristics and standardised differences of trial-eligible patients after applying trial criteria included in propensity-score—weighted analysis before and after weighting using operational definition of dual user

| Characteristic | Before weighting | | After weighting | | |
|---|---|---|---|---|---|
| | **Dual therapy** N=19 931 | **ACEi** N=392 475 | **Dual therapy** N=399 678 | **ACEi** N=412 551 | **SMD** |
| Oral anticoagulant agent | 1147 (5.8) | 27971 (7.1) | 29554 (7.4) | 29134 (7.1) | 0.013 |
| Antiplatelet agent | 1442 (7.2) | 31143 (7.9) | 33168 (8.3) | 32632 (7.9) | 0.014 |
| Aspirin | 7809 (39.2) | 135630 (34.6) | 135945 (34.0) | 143522 (34.8) | 0.016 |
| Beta-blocker | 6492 (32.6) | 120585 (30.7) | 120981 (30.3) | 127133 (30.8) | 0.012 |
| Calcium-channel blocker | 7917 (39.7) | 130355 (33.2) | 129784 (32.5) | 138351 (33.5) | 0.023 |
| Digoxin | 557 (2.8) | 13976 (3.6) | 11459 (2.9) | 14664 (3.6) | 0.039 |
| Diuretics | 9519 (47.8) | 150598 (38.4) | 156134 (39.1) | 160298 (38.9) | 0.004 |
| Diabetic treatment | 7077 (35.5) | 101350 (25.8) | 99904 (25.0) | 108543 (26.3) | 0.030 |
| Nitrates | 1730 (8.7) | 36904 (9.4) | 37505 (9.4) | 38650 (9.4) | 0.001 |
| Statins | 11508 (57.7) | 210608 (53.7) | 208064 (52.1) | 222277 (53.9) | 0.036 |
| **Time-related variables – mean (SD)** | | | | | |
| Time since trial-eligible period | 315.1 (780.3) | 320.8 (838.4) | 328.9 (854.8) | 320.6 (835.8) | 0.010 |
| Calendar year | 2008 (4.3) | 2010 (5.3) | 2010 (4.7) | 2010 (5.3) | 0.027 |
| **Healthcare utilisation[f] – mean (SD)** | | | | | |
| Number of GP appointments | 0.8 (3.5) | 7.6 (25.4) | 1.9 (5.5) | 7.4 (25.0) | 0.100 |
| Number of hospital admissions | 0.3 (2.2) | 1.0 (3.4) | 1.3 (5.1) | 1.0 (3.4) | 0.016 |
| **Index of multiple deprivation – no. (%)** | | | | | |
| 1 (least) | 3783 (19.0) | 75076 (19.1) | 76720 (19.2) | 78912 (19.1) | 0.002 |
| 2 | 4060 (20.4) | 80031 (20.4) | 82442 (20.6) | 84099 (20.4) | 0.006 |
| 3 | 4015 (20.1) | 77078 (19.6) | 78788 (19.7) | 81142 (19.7) | 0.001 |
| 4 | 4137 (20.8) | 80891 (20.6) | 82437 (20.6) | 85050 (20.6) | 0.000 |
| 5 (most) | 3936 (19.8) | 79399 (20.2) | 79292 (19.8) | 83348 (20.2) | 0.009 |

N= number of patients; SD= standard deviation; no. (%)=number (percent); SD=standard deviation; SMD=standardised mean difference; BP= blood pressure; CAD=coronary artery disease; PAD=peripheral artery disease

Post-weighting N displays weighted distribution of number of patients in the two exposure groups.

Propensity-score weights are unstabilized inverse probability weights.

Number of GP and hospital admissions were transformed to the log scale.

**Supplementary Table S5.** Baseline characteristics and standardised differences of trial-eligible patients after applying trial criteria included in propensity-score—weighted analysis before and after weighting using operational definition of dual user

| Characteristic | Before weighting | | After weighting | | |
|---|---|---|---|---|---|
| | **Dual therapy** N=19 931 | **ACEi** N=392 475 | **Dual therapy** N=399 678 | **ACEi** N=412 551 | **SMD** |

One third of ONTARGET participants received both ramipril plus telmisartan.

[a] Includes diagnosis of: MI at least 2 days prior, angina at least 30 days prior, angioplasty at least 30 days prior, CABG at least 4 years prior

[b] Includes diagnosis of: stroke/TIA

[c] Includes diagnosis of: limb bypass surgery, limb/foot amputation, intermittent claudication

[d] Includes DM with: retinopathy, neuropathy, chronic kidney disease, proteinuria or other complication

[e] Within 3 months prior to eligible start date. Antiplatelet agent= clopidogrel/ticlopidine

[f] Within 6 months prior to eligible start date.
