## Supplementary Table S6 for "Cardiorenal effects of dual blockade with Angiotensin-converting enzyme inhibitors and Angiotensin receptor blockers in people with CKD: analysis of routinely collected data with emulation of a reference trial (ONTARGET)"

**Supplementary Table S6.** Baseline characteristics of trial-eligible patients after applying trial criteria included in propensity-score—weighted analysis using primary definition and alternate definitions of dual users

| **Characteristics** | **Primary definition** N=19 931 | **Alternate 1 definition** N=13,095 | **Alternate 2 definition** N=20,502 |
|---|---|---|---|
| **Age (year) – mean (SD)** | 70.4 (8.8) | 70.5 (8.7) | 70.1 (8.6) |
| **Systolic BP (mmHg) – mean (SD)** | 147.7 (20.0) | 150.6 (22.3) | 148.5 (20.1) |
| **Diastolic BP (mmHg) – mean (SD)** | 79.7 (10.8) | 80.0 (11.4) | 79.9 (10.7) |
| **Body-mass index – mean (SD)** | 29.7 (5.9) | 29.9 (6.0) | 29.7 (5.8) |
| **Creatinine (µmol/l) – mean (SD)** | 96.4 (28.9) | 99.8 (32.3) | 96.8 (28.6) |
| **Potassium (mmol/l) – mean (SD)** | 4.5 (0.5) | 4.4 (0.5) | 4.4 (0.5) |
| **Female sex – no. (%)** | 10388 (52.1) | 6512 (49.7) | 10652 (52.0) |
| **Ethnic group – no. (%)** | | | |
| Black | 851 (4.3) | 720 (5.5) | 881 (4.2) |
| South Asian | 1565 (7.9) | 1093 (8.4) | 1634 (8.0) |
| White | 17186 (86.2) | 11023 (84.2) | 17650 (86.1) |
| Other | 329 (1.7) | 259 (2.0) | 337 (1.6) |
| **Clinical history – no. (%)** | | | |
| CAD[a] | 12296 (61.7) | 11253 (85.9) | 12264 (59.8) |
| Myocardial infarction | 3965 (19.9) | 4094 (31.3) | 3665 (17.9) |
| Cerebrovascular disease[b] | 2149 (10.8) | 1143 (8.7) | 2060 (10.1) |
| PAD[c] | 1930 (9.7) | 1454 (11.1) | 1917 (9.4) |
| Diabetes | 13462 (67.5) | 8704 (66.5) | 13592 (66.3) |
| High-risk diabetes[d] | 12574 (63.1) | 7356 (56.1) | 12962 (63.2) |
| **Smoking status – no. (%)** | | | |
| Non-smoker | 6040 (30.3) | 3870 (29.6) | 9031 (45.3) |
| Current smoker | 4860 (24.4) | 3226 (24.6) | 5999 (45.8) |
| Past smoker | 9031 (45.3) | 5046 (24.6) | 9027 (44.0) |
| **Alcohol status – no. (%)** | | | |
| Non-drinker | 3711 (18.6) | 2567 (19.6) | 3949 (19.3) |
| Current drinker | 11578 (58.1) | 7538 (57.6) | 11834 (57.7) |
| Past drinker | 2647 (13.3) | 1811 (13.8) | 2527 (12.3) |
| Missing | 1995 (10.0) | 1179 (9.0) | 2192 (10.7) |
| **Medication[e] – no. (%)** | | | |
| Alpha-blocker | 2817 (14.1) | 2669 (20.4) | 2905 (14.2) |

**Supplementary Table S6.** Baseline characteristics of trial-eligible patients after applying trial criteria included in propensity-score—weighted analysis using primary definition and alternate definitions of dual users

| **Characteristics** | **Primary definition** N=19 931 | **Alternate 1 definition** N=13,095 | **Alternate 2 definition** N=20,502 |
|---|---|---|---|
| Oral anticoagulant agent | 1147 (5.8) | 1077 (8.2) | 1113 (5.4) |
| Antiplatelet agent | 1442 (7.2) | 1175 (9.0) | 1591 (7.8) |
| Aspirin | 7809 (39.2) | 5699 (43.5) | 8391 (40.9) |
| Beta-blocker | 6492 (32.6) | 4944 (37.8) | 6918 (33.7) |
| Calcium-channel blocker | 7917 (39.7) | 5812 (44.4) | 8312 (40.5) |
| Digoxin | 557 (2.8) | 593 (4.5) | 549 (2.7) |
| Diuretics | 9519 (47.8) | 7064 (53.9) | 10186 (49.7) |
| Diabetic treatment | 7077 (35.5) | 5050 (38.6) | 7379 (36.0) |
| Nitrates | 1730 (8.7) | 1469 (11.2) | 1888 (9.2) |
| Statins | 11508 (57.7) | 8004 (61.1) | 12011 (58.6) |
| **Healthcare utilisation[f] – mean (SD)** | | | |
| Number of GP appointments | 0.8 (3.5) | 6.0 (24.9) | 3.4 (6.9) |
| Number of hospital admissions | 0.3 (2.2) | 3.0 (12.1) | 1.2 (4.8) |
| **Index of multiple deprivation – no. (%)** | | | |
| 1 (least) | 3783 (19.0) | 2205 (16.8) | 3972 (19.4) |
| 2 | 4060 (20.4) | 2513 (19.2) | 4216 (20.6) |
| 3 | 4015 (20.1) | 2643 (20.2) | 4161 (20.3) |
| 4 | 4137 (20.8) | 2899 (22.1) | 4251 (20.7) |
| 5 (most) | 3936 (19.8) | 2835 (21.7) | 3902 (19.0) |

One third of ONTARGET participants received both ramipril plus telmisartan.

[a] Includes diagnosis of: MI at least 2 days prior, angina at least 30 days prior, angioplasty at least 30 days prior, CABG at least 4 years prior

[b] Includes diagnosis of: stroke/TIA

[c] Includes diagnosis of: limb bypass surgery, limb/foot amputation, intermittent claudication

[d] Includes DM with: retinopathy, neuropathy, chronic kidney disease, proteinuria or other complication

[e] Within 3 months prior to eligible start date. Antiplatelet agent= clopidogrel/ticlopidine

[f] Within 6 months prior to eligible start date.
